# Exposure-response analysis of the association of maternal smoking and use of electronic cigarettes (vaping) in relation to preterm birth and small-for-gestational-age in a national US sample, 2016-2018

**DOI:** 10.1101/2021.03.01.21252530

**Authors:** Xi Wang, Nora L Lee, Igor Burstyn

**Author notes:** **Corresponding Author:** Xi Wang, PhD, Mailing address: Children’s Hospital of Philadelphia PolicyLab, 2716 South Street, 10th Floor, Philadelphia, PA 19146. **Disclosure of Interest:** The authors report no conflict of interest. **Funding Source:** This research did not receive any specific grant from funding agencies in the public, commercial, or not-for-profit sectors.

## Abstract

We aimed to estimate exposure-response associations between smoking or vaping, and preterm and small-for-gestational age (SGA) births. We included 99,201 mothers who delivered live singletons in 2016-2018 from the Pregnancy Risk Assessment Monitoring System. We created exposure categories based on participants’ self-reported average number of cigarettes smoked per day and vaping frequency. Dual users in late pregnancy were a heterogeneous group: 36% heavily smoked and occasionally vaped; 29% lightly smoked and frequently vaped; 19% lightly smoked and frequently vaped; and 15% both heavily smoked and frequently vaped. While dual users who heavily smoked and occasionally vaped had the highest adjusted OR for SGA (3.4, 95% CI 1.7-6.6), all the dual users were on average at about twice the odds of having SGA than non-users. While the risks of preterm birth were higher among sole light smokers (adjusted OR 1.3, 95% CI 1.1-1.5) and sole heavy smokers (adjusted OR 1.4. 95 CI 1.2-1.8) than non-users, the adjusted odds of preterm births for dual users were not noticeably higher than those of non-users, unless they were also heavy smokers. Excess of preterm births among heavy vapers was suggested. Among younger non-Hispanic white women (where vaping is most common), only excess risk of SGA, not preterm, with vaping was apparent. Relative to non-users, both smoking and vaping during pregnancy appear to increase risk of SGA, but excess risk of preterm births appears to be primarily attributable to smoking alone. Higher levels of exposure tended to confer more risk.

**Highlights:**

- No observable change in prevalence of vaping during pregnancy from 2016 to 2018
- Both smoking and vaping during pregnancy appear to increase risk of SGA births
- Excess risk of preterm births appears to be primarily attributable to smoking alone

## 1. INTRODUCTION

Smoking during pregnancy is a major public health concern in the US, with 8.4% of women reporting smoking before pregnancy and 6.5% during pregnancy in 2018.^1^ Maternal smoking during pregnancy is consistently associated with adverse birth outcomes such as preterm birth and low birth weight, as well as fetal and infant mortality.^2^ Since its emergence as a perceived safer alternative to smoking, the US experienced a surge in vaping or use of electronic cigarettes (e-cigarettes), especially among adolescents and young adults.^3^ Women who smoke and are pregnant or planning to become pregnant may be more likely to consider using e-cigarettes as an alternative to smoking.^4^

There is insufficient knowledge about the effect of vaping on pregnancy outcomes. Cardenas et al. (2019), in a systematic review of 96 studies published in 2007-2018, did not find any human studies that evaluated the effects of e-cigarette use during pregnancy on reproductive outcomes. Most of the 96 studies used animals or tissue models.^5^ More recently, Glover and Phillips (2020) reviewed the literature on smoke-free nicotine and tobacco product exposure (nicotine replacement therapies, Swedish snus, Alaskan iq’mik, and vaping) and birth outcomes, concluding that “smoke-free product use during pregnancy probably increases the risk of some negative birth outcomes, but that any effect is less than that from smoking”.^6^ Confounding by socioeconomic factors is one of the unresolved issues in the literature, given the known association of vaping with established correlates of adverse pregnancy outcomes in the US (e.g., race, lower education, younger age).

Moreover, when evaluating the pregnancy outcomes associated with vaping, few studies have appropriately accounted for vapers also smoking combustible cigarettes (dual users). A 2013–2014 US national study found that most (79%) of the e-cigarette users smoked cigarettes contemporanously.^7^ Specific to pregnant women, a 2016 US nationally representative study suggested that a majority (64%) of pregnant women who vaped also concurrently smoked.^8^ Therefore, it is crucial for studies on vaping and pregnancy outcomes to capture exposure in women who use both products, given the well-established evidence on the adverse effects of traditional smoking on pregnancy, and because it is not possible to assert without any evidence that the effect of dual use will merely be the sum of the effects of vaping and smoking. Unfortunately, published analysis of the most comprehensive dataset able to inform the issue in the US excluded dual users.^9^ Importantly, dual users are not a homogeneous group when it comes to relative exposures from smoking versus vaping, because they include both habitual heavy smokers who occasionally vape and vapers who only occasionally smoke. Due to the well-known dose-response relationship between smoking and the risks of fetal and neonatal morbidities, it is reasonable to examine whether the intensity of smoking and vaping relates to pregnancy outcomes in a manner consistent with exposure-response, one of the elements in evaluating causation.

We investigated the effect of vaping during pregnancy on neonatal outcomes in a US nationally representative sample from 2016-2018 with the primary aim of assessing exposure-response associations while accounting for concurrent smoking and vaping.

## 2. METHODS

### 2.1 Sample and data source

We used data from the 2016-2018 Pregnancy Risk Assessment Monitoring System (PRAMS), a surveillance project of the US Centers for Disease Control and Prevention (CDC) with state health departments. Details of its design and methodology are described elsewhere.^10^ Briefly, PRAMS samples women who have had a recent live birth from the state’s birth certificates, which currently covers about 83% of all US births. We restricted our sample to 2016-2018 PRAMS participants who had singleton births (excluding 3,806 multiple births and 2,648 births with missing plurality) and have complete information on smoking and vaping before and during pregnancy (excluding 2,455 mothers with missing values). Consequently, 99,201 mother-birth dyads were included (weighted sample of 5,333,904). All PRAMS data were de-identified, and the analysis was deemed by the Institutional Review Board of Drexel University as being exempt from review.

### 2.2 Sole and dual uses of traditional and e-cigarettes

Starting in 2016, questions on mother’s use of e-cigarettes were added for all PRAMS states. In the PRAMS core questionnaire, e-cigarettes and other electronic nicotine products (such as vape pens, e-hookahs, hookah pens, e-cigars, e-pipes) are defined as battery-powered devices that use nicotine liquid rather than tobacco leaves, and produce vapor instead of smoke. Participants were asked the frequency of using e-cigarettes or other electronic nicotine products in two separate time periods: during the 3 months before their pregnancy, and during the last 3 months of pregnancy (i.e., late pregnancy). Daily average quantities of cigarettes smoked was also queried for those two time periods.

We created four exposure groups based on participants’ reported smoking and vaping during each of the two time periods: non-users, sole smokers, sole vapers, and dual users. To further refine the dose of nicotine and combustible products, we created nine categories of exposure based on participants’ self-reported average number of cigarettes smoked per day and vaping frequency (see **Table 2** for definitions of the nine exposure categories). Heavy smokers were defined as those who smoked on average ≥6 combustible cigarettes per day. Frequent vapers were defined as those who vaped on average ≥ once per day. Dual users were then divided into four subgroups: a) heavy smoker and frequent vaper; b) heavy smoker and occasional vaper; c) light smoker and frequent vaper; and d) light smoker and occasional vaper.

### 2.3 Pregnancy Outcomes

We chose to investigate risk of preterm birth and small-for-gestational-age (SGA) births based on prior literature on maternal exposure to nicotine,^11,12^ their availability in nationwide PRAMS data, and the sample size. Gestational age at delivery in completed weeks is based on the clinician’s best obstetric estimate. In PRAMS, SGA was defined as births with weight lower than the tenth percentile of the population, which was calculated from the 2006 national natality file for singleton births for each gestational age, race/ethnicity, and infant sex combination.

### 2.4 Covariates

We selected potential confounders a priori, including birth year, mother’s demographic characteristics (age, educational level, race/ethnicity, marital status), parity (number of previous live births), previous preterm delivery, adequacy of prenatal care, and maternal pre-pregnancy body mass index (BMI), as consistent with our previous work on this topic and available data.^8^ We calculated an index of adequacy of visits based on the initiation time and number of prenatal care visits, and categorized women as receiving intensive, adequate, intermediate, or inadequate prenatal care.^13^

### 2.5 Statistical Methods

We described the proportions of mothers using tobacco cigarettes and/or e-cigarettes before and during late pregnancy, in the full sample and stratified by their sociodemographic and clinical characteristics. Multivariable logistic models were fitted to estimate the associations between maternal smoking and vaping in late pregnancy and pregnancy outcomes in terms of adjusted odds ratios (aOR) and 95% confidence intervals (CI). In addition to the covariates listed above, we adjusted for pre-pregnancy (three months) smoking and vaping, categorized as non-users, sole smokers, sole vapers, and dual users. We also estimated the marginal predicted probabilities (PP) for pregnancy outcomes by maternal smoking and vaping. We applied statistical weighting schemes to account for different sampling rates in different strata and nonresponse. To examine whether the effect estimates (aORs) were similar in subgroups as those in the full sample, we conducted two sensitivity analyses in which we only included young mothers (aged <25 years old at the time of delivery) and non-Hispanic mothers, separately, because these two groups had the highest prevalence of vaping. Analyses were conducted using SAS 9.4 and R.

## 3. RESULTS

### 3.1 Reported prevalence of smoking and vaping

**Table 1** presents the frequencies and weighted percentages of our study sample by their reported smoking and vaping before and during late pregnancy. There is no noticeable annual change in smoking and vaping rate from 2016-2018. The proportions of the sole smokers, sole vapers, and dual users in mothers who had live births in 2018 were like those in 2016 and 2017. Therefore, we combined the PRAMS 2016-2018 data, and adjusted for childbirth year to account for any residual temporal confounding. Among mothers who had singleton live births in the 2016-2018 PRAMS, 14.3% exclusively smoked cigarettes (“sole smokers”), 1.0% exclusively used e-cigarettes (“sole vapers”), and 2.6% used both products (“dual users”) in the three months before pregnancy. Fewer used either product in late pregnancy, with 7.1% sole smokers, 0.4% sole vapers, and 0.7% dual users in the last three months of pregnancy.

**Table 1.** Prevalence of smoking and vaping before and during pregnancy by the year of childbirth, in 2016-2018 PRAMS singleton births

|  | 2016 |  | 2017 |  | 2018 |  | 2016-2018 |  |
| --- | --- | --- | --- | --- | --- | --- | --- | --- |
|  | Unweight<br>ed N | Weighte<br>d<br>column<br>%<br>(95%<br>Confiden<br>ce<br>Interval) | Unweight<br>ed N | Weighte<br>d<br>column<br>%<br>(95%<br>Confiden<br>ce<br>Interval) | Unweight<br>ed N | Weighte<br>d<br>column<br>%<br>(95%<br>Confiden<br>ce<br>Interval) | Unweight<br>ed N | Weighte<br>d<br>column<br>%<br>(95%<br>Confiden<br>ce<br>Interval) |
| **All** | 32,791 |  | 35,530 |  | 30,880 |  | 99,201 |  |
| **Pre-pregnancy, in the 3 months before pregnancy (covariate)** |  |  |  |  |  |  |  |  |
| **Sole**<br>**smoke**<br>**rs** | 5,141 | 14.1<br>(13.5-14.7) | 5,838 | 15.0<br>(14.4-15.6) | 4,683 | 13.8<br>(13.2-14.3) | 15,662 | 14.3<br>(14.0-14.7) |
| **Sole**<br>**vapers** | 285 | 0.8<br>(0.7-1.0) | 332 | 1.1<br>(0.9-1.2) | 351 | 1.1<br>(0.9-1.2) | 968 | 1.0<br>(0.9-1.1) |
| **Dual**<br>**users** | 1,000 | 2.8<br>(2.5-3.1) | 892 | 2.6<br>(2.4-2.9) | 737 | 2.3<br>(2.1-2.6) | 2,629 | 2.6<br>(2.4-2.8) |
| **Non-**<br>**users** | 26,365 | 82.2<br>(81.6-82.9) | 28,468 | 81.3<br>(80.7-81.9) | 25,109 | 82.8<br>(82.2-83.5) | 79,942 | 82.1<br>(81.7-82.5) |
| **Late pregnancy, in the last 3 months of pregnancy (primary exposure)** |  |  |  |  |  |  |  |  |
| **Sole**<br>**smoke**<br>**rs** | 2,674 | 6.9<br>(6.5-7.4) | 3,020 | 7.5<br>(7.0-7.9) | 2,470 | 6.9<br>(6.5-7.3) | 8,164 | 7.1<br>(6.9-7.4) |
| **Sole**<br>**vapers** | 133 | 0.4<br>(0.3-0.5) | 141 | 0.4<br>(0.3-0.5) | 132 | 0.4<br>(0.3-0.5) | 406 | 0.4<br>(0.3-0.5) |
| **Dual**<br>**users** | 267 | 0.7<br>(0.5-0.8) | 246 | 0.7<br>(0.5-0.8) | 210 | 0.7<br>(0.5-0.8) | 723 | 0.7<br>(0.6-0.8) |
| **Non-**<br>**users** | 29,717 | 92.0<br>(91.6-92.5) | 32,123 | 91.4<br>(91.0-91.9) | 28,068 | 92.0<br>(91.6-92.5) | 89,908 | 91.8<br>(91.6-92.1) |

**Table 2** presents the proportions of sole smokers, sole vapers, and dual users in each group of mothers categorized by their sociodemographic and clinical characteristics. Proportions of sole vapers and dual users were higher in the groups of younger mothers, mothers with low educational attainment, non-Hispanic white mothers, not-married mothers, pre-pregnancy underweight mothers, and mothers receiving inadequate prenatal care. For example, 2.6% of mothers under 20 years old were sole vapers or dual users in late pregnancy, while the proportion was 0.6% among mothers 35 years and older.

**Table 2.**
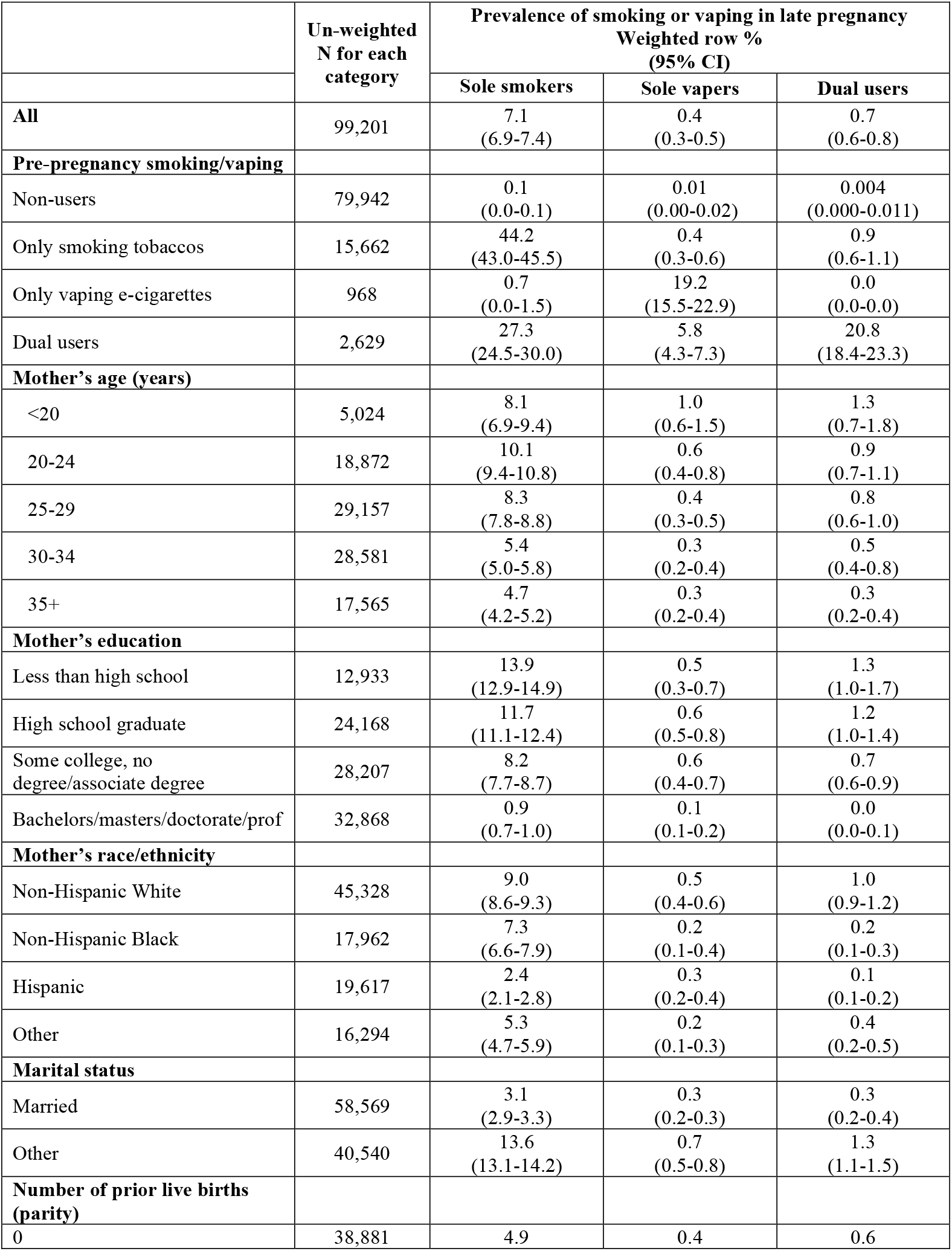
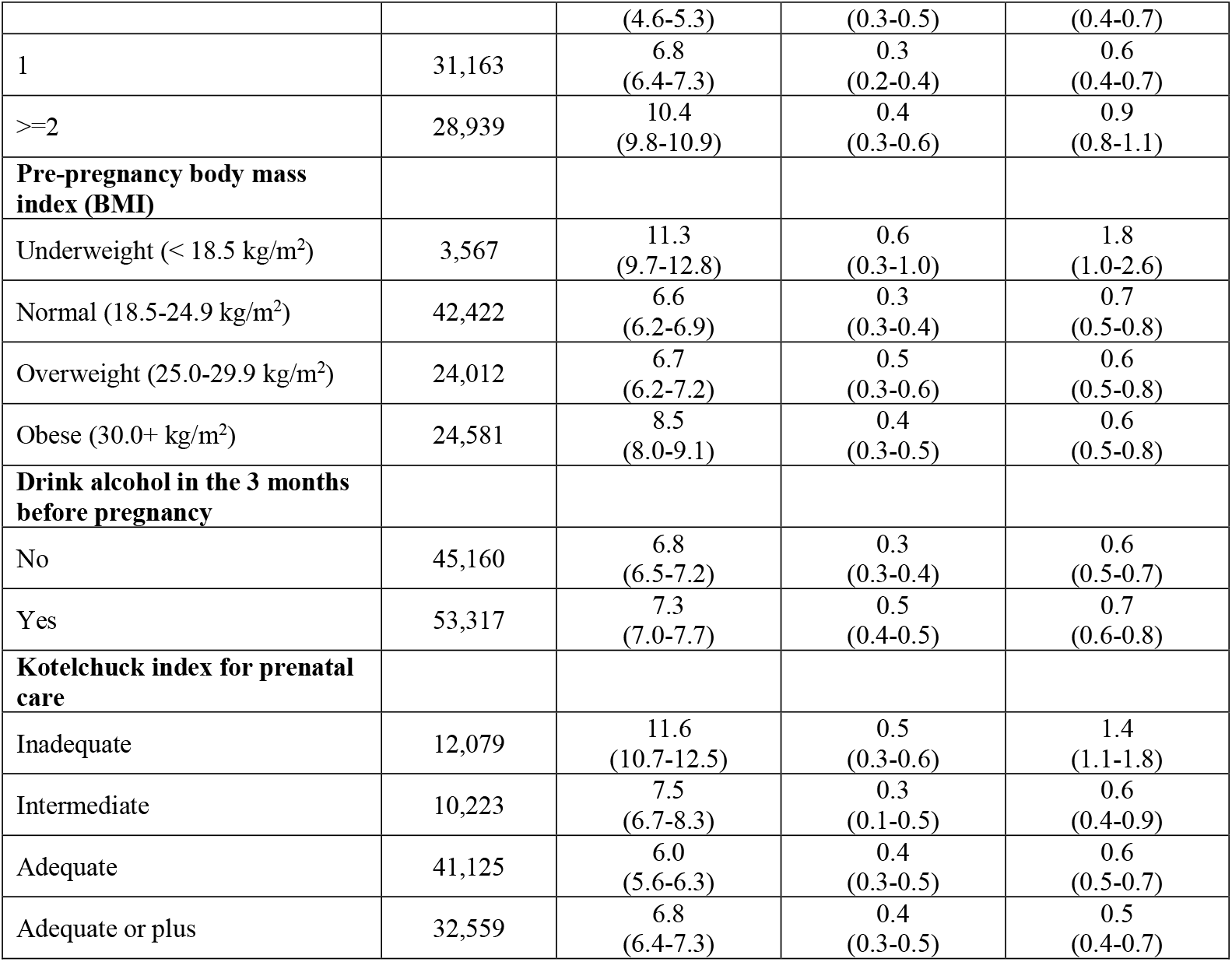
Prevalence of vaping (sole vapers or dual users) during late pregnancy by maternal characteristics, in 2016-2018 PRAMS singleton births

**Table 3** presents the definitions and prevalence of mothers grouped by their smoking amount and vaping frequency in late pregnancy. The nine groups were sorted based on our assumption on their level of exposure to cigarette combustion by-products and nicotine, from the lowest to the highest for each. Among the 8% of pregnant women who used either combustible or e-cigarettes, sole light smokers were the most common, with 4.2% of pregnant women reporting exclusively smoking on average 1-5 cigarettes per day in late pregnancy. Dual users in late pregnancy were a heterogeneous group regarding smoking amount and vaping frequency: 36% heavily smoked (≥6 cigarettes per day) and occasionally vaped (< once per day); 29.3% lightly smoked (1-5 cigarettes per day) and frequently vaped (≥ once per day); 19.4% lightly smoked and frequently vaped; and 15.2% both heavily smoked and frequently vaped.

**Table 3.**
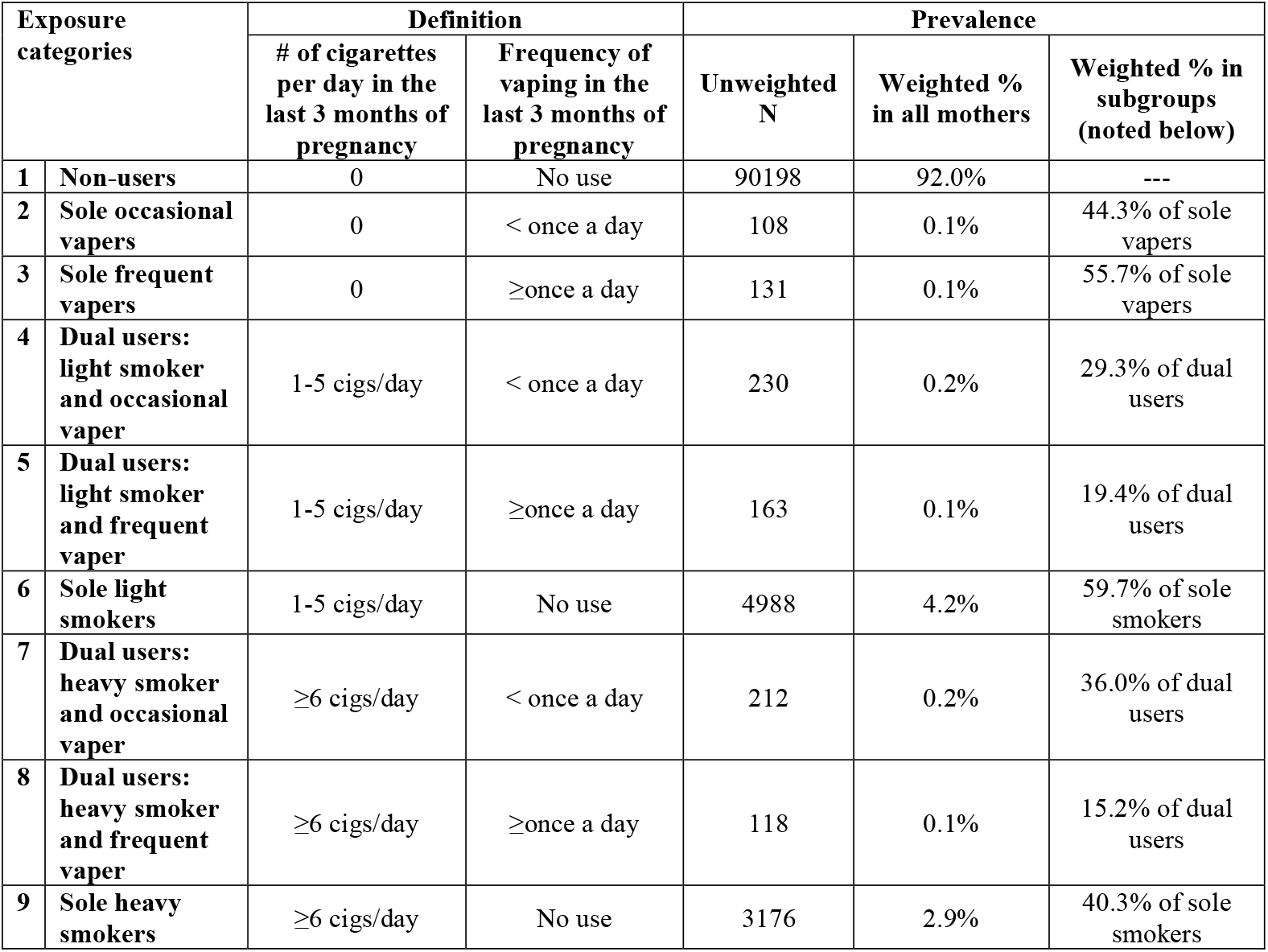
Definition and prevalence of exposure categories of smoking and vaping in late pregnancy, among 2016-2018 PRAMS singleton births.

### 3.2 Pregnancy outcomes associated with the exposure categories of smoking and vaping in late pregnancy

**Tables 4** and **5** present the raw incidences of preterm births and SGA births, respectively, by reported maternal smoking amount and vaping frequency in late pregnancy and the adjusted OR after controlling for sociodemographic and obstetric characteristics. Considering pre-pregnancy behavior as a plausible confounder, we fitted two models without (Model a) and with (Model b) adjusting for pre-pregnancy smoking and/or vaping for each outcome.

**Table 4.** Incidences and adjusted odds ratios (ORs) and 95% confidence interval (CI) for <u>preterm births</u> associated with dose of smoking and vaping in late pregnancy, among 2016-2018 PRAMS singleton births.

| Dose of smoking and vaping in late pregnancy |  | Unweighted N | PRETERM BIRTH |  |  |
| --- | --- | --- | --- | --- | --- |
|  |  |  | Incidence % | Adjusted OR (95% CI)* |  |
|  |  |  |  | Model a | Model b |
| 1 | Non-users | 90198 | 7.6 | Reference | Reference |
| 2 | Sole occasional vapers | 108 | 5.1 | 0.8 (0.4-1.7) | 0.8 (0.3-1.7) |
| 3 | Sole frequent vapers | 131 | 10.0 | 1.3 (0.7-2.3) | 1.2 (0.7-2.3) |
| 4 | Dual users: light smoker and occasional vaper | 230 | 5.5 | 0.7 (0.4-1.1) | 0.6 (0.3-1.1) |
| 5 | Dual users: light smoker and frequent vaper | 163 | 8.7 | 1.2 (0.6-2.4) | 1.2 (0.6-2.4) |
| 6 | Sole light smokers | 4988 | 11.1 | 1.3 (1.2-1.5) | 1.3 (1.1-1.5) |
| 7 | Dual users: heavy smoker and occasional vaper | 212 | 11.3 | 1.6 (0.8-2.9) | 1.5 (0.8-2.9) |
| 8 | Dual users: heavy smoker and frequent vaper | 118 | 8.5 | 1.4 (0.8-2.6) | 1.4 (0.8-2.6) |
| 9 | Sole heavy smokers | 3176 | 11.7 | 1.5 (1.3-1.8) | 1.4 (1.2-1.8) |

\* **Model a** adjusted for mother's age, education level, race/ethnicity, marital status, previous preterm history, plurality (number of previous live births), Kotelchuck index of prenatal care, pre-pregnancy BMI (underweight, normal, overweight, or obesity), drinking alcohol before pregnancy, and birth year. **Model b** further adjusted for pre-pregnancy (in the 3 months before pregnancy) smoking/vaping (categorized as non-users, sole smokers, sole vapers, and dual users).

**Table 5.** Incidences and adjusted odds ratios (ORs) and 95% confidence interval (CI) for <u>small-for-gestational-age births</u> associated with dose of smoking and vaping in late pregnancy, among 2016-2018 PRAMS singleton births.

| Dose of smoking and vaping<br>in late pregnancy |  | Unweighted<br>N | SMALL-FOR-GESTATIONAL-AGE (SGA) BIRTH |  |  |
| --- | --- | --- | --- | --- | --- |
|  |  |  | Incidence % | Adjusted OR* |  |
|  |  |  |  | Model a | Model b |
| 1 | Non-users | 90198 | 8.9 | Reference | Reference |
| 2 | Sole occasional vapers | 108 | 15.1 | 1.4 (0.5-4.0) | 1.4 (0.5-3.9) |
| 3 | Sole frequent vapers | 131 | 10.1 | 1.1 (0.5-2.5) | 1.1 (0.5-2.5) |
| 4 | Dual users: light smoker<br>and occasional vaper | 230 | 18.7 | 2.0 (1.2-3.3) | 2.0 (1.1-3.4) |
| 5 | Dual users: light smoker<br>and frequent vaper | 163 | 27.0 | 3.4 (1.8-6.4) | 3.4 (1.7-6.6) |
| 6 | Sole light smokers | 4988 | 18.1 | 2.2 (1.9-2.5) | 2.1 (1.8-2.5) |
| 7 | Dual users: heavy smoker<br>and occasional vaper | 212 | 25.3 | 3.4 (2.1-5.5) | 3.4 (2.0-5.7) |
| 8 | Dual users: heavy smoker<br>and frequent vaper | 118 | 20.8 | 2.9 (1.3-6.5) | 3.1 (1.3-7.1) |
| 9 | Sole heavy smokers | 3176 | 21.5 | 2.9 (2.5-3.3) | 2.8 (2.3-3.3) |

\* **Model a** adjusted for mother's age, education level, race/ethnicity, marital status, previous preterm history, plurality (number of previous live births), Kotelchuck index of prenatal care, pre-pregnancy BMI (underweight, normal, overweight, or obesity), drinking alcohol before pregnancy, and birth year. **Model b** further adjusted for pre-pregnancy (in the 3 months before pregnancy) smoking/vaping (categorized as non-users, sole smokers, sole vapers, and dual users).

In **Table 4**, we observed that the late-pregnancy sole smokers had higher odds of preterm birth than non-user, with some evidence of exposure-response: aOR of 1.3 (95% CI 1.1-1.5) for sole light smokers increased to aOR of 1.4 (95% CI 1.2-1.8) among sole heavy smokers. Dual users who were occasional vapers and light smokers, as well as occasional vapers, had similar odds of preterm birth to non-users, with point estimates of aOR just under 1.0. Notably, among frequent vapers the incidence of preterm birth was like that of sole smokers (light and heavy), at 10.0%, with an aOR of 1.2 (95% CI 0.7-2.3) that was lower on average yet comparable to sole smokers. The association with dual use that included frequent vaping and light smoking was like that of frequent vaping alone: aOR 1.2 (95% CI 0.6-2.4). On average, elevated yet imprecise estimates of aOR among dual users appear to be driven by co-occurrence of heaving smoking.

In **Table 5**, we observed that the late-pregnancy sole smokers (light and heavy) had higher odds of SGA births than non-users, with associations that support higher risk with higher exposure. Two subgroups of dual users -- dual users who lightly smoked and frequently vaped, and dual users who heavily smoked and occasionally vaped -- had the highest incidences of SGA births at 27.0% and 25.3%, respectively.

While dual users who heavily smoked and occasionally vaped had the highest aOR for SGA (3.4, 95% CI 1.7-6.6), all the dual-user subgroups were associated with, on average, at least twice the odds of having SGA births regardless of their smoking amount and vaping frequency. There was no evidence of excess SGA risk among sole vapers and no evidence of the anticipated exposure-response.

We saw no noticeable effect of adjustment for pre-pregnancy smoking and vaping (Model b) on the patterns described above (**Tables 4** and **5**). We explored potential effect modification by pre-pregnancy smoking and vaping. **Figures 1** and **2** present the model-based probabilities of pregnancy outcomes by late-pregnancy smoking and vaping dose categories, stratified by pre-pregnancy smoking and vaping. We found no evidence for differential effects: the associations between late-pregnancy smoking and vaping and pregnancy outcomes were similar in different strata of women grouped by their pre-pregnancy smoking/vaping status.

**Figure 1.**
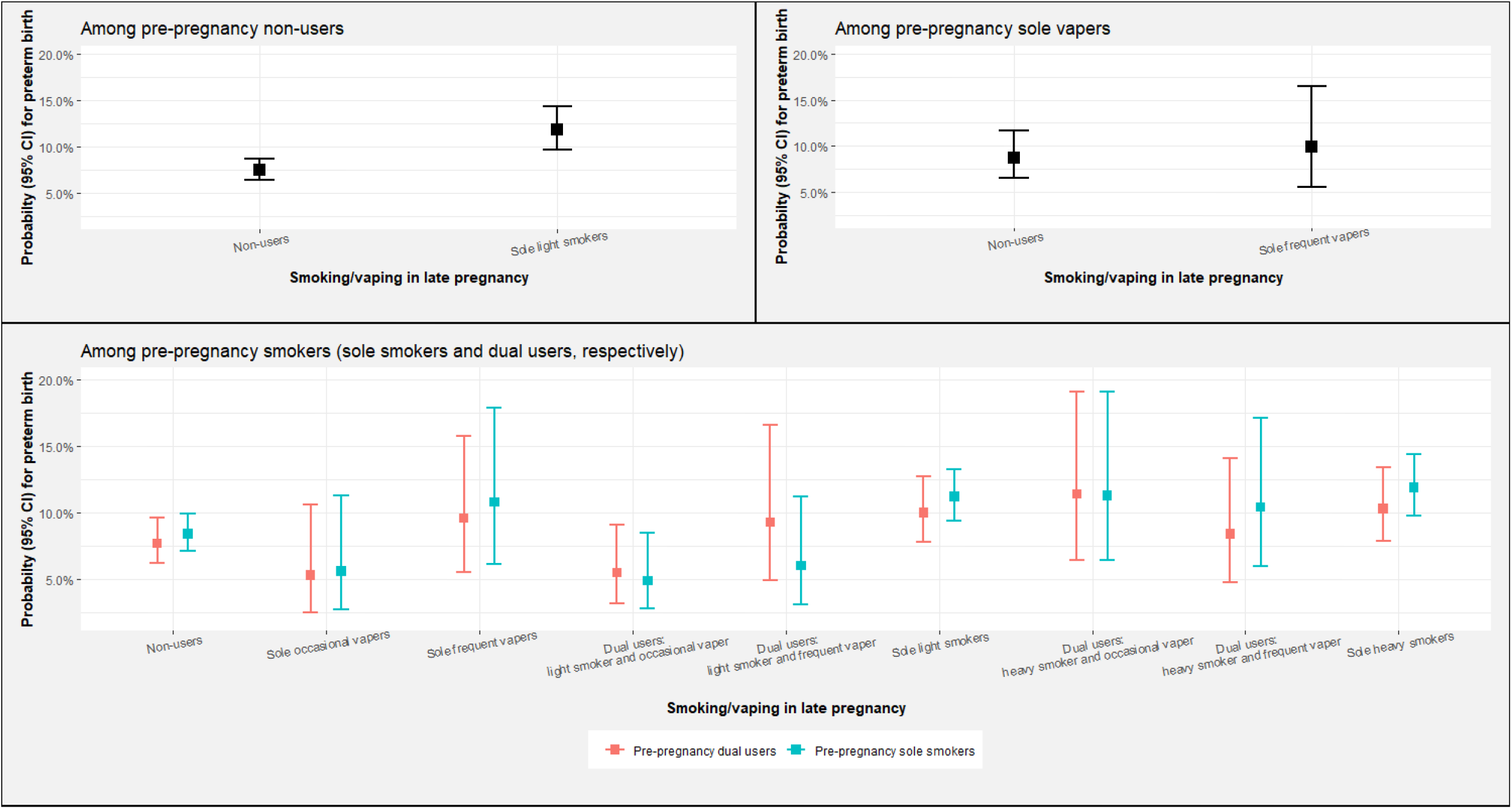
Predicted probabilities for preterm birth by maternal smoking or vaping in late pregnancy, stratified by smoking or vaping before pregnancy • This figure presents the model-based predicted probabilities (PP) for preterm birth by maternal smoking and vaping in late pregnancy, stratified by maternal smoking and vaping before conception. Covariates in the models included mother’s age, education level, race/ethnicity, marital status, previous preterm history, plurality (number of previous live births), Kotelchuck index of prenatal care, pre-pregnancy BMI (underweight, normal, overweight, or obesity), drinking alcohol before pregnancy, and birth year. Groups with less than 20 mothers in the sample were not included in the figure (see **Appendix Table A3** for cell sizes of included and excluded groups).

**Figure 2.**
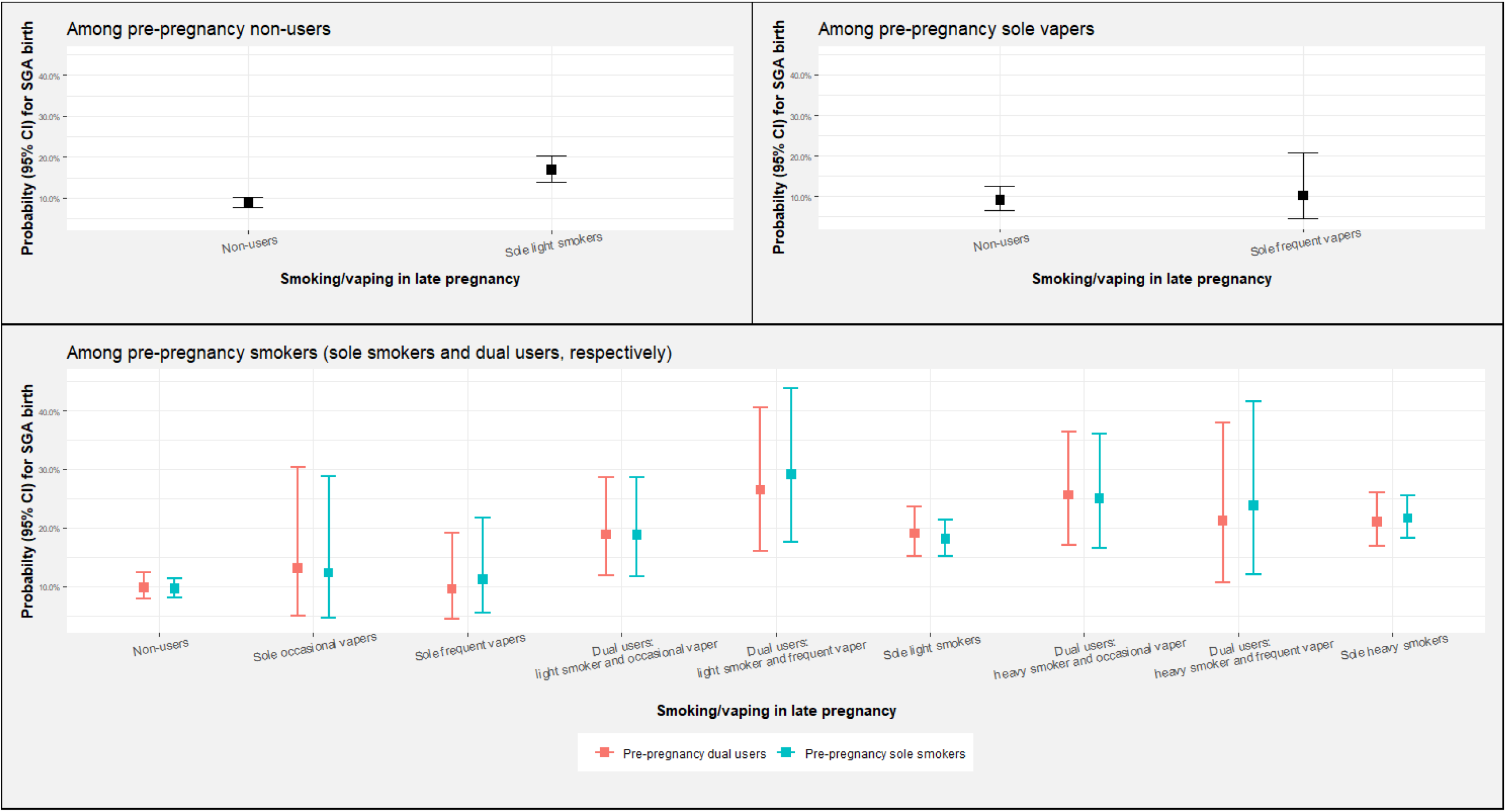
Predicted probabilities for small-for-gestational-age (SGA) birth by maternal smoking or vaping in late pregnancy, stratified by smoking or vaping before pregnancy • This figure presents the model-based predicted probabilities (PP) for small-for-gestational-age (SGA) birth by maternal smoking and vaping in late pregnancy, stratified by maternal smoking and vaping before conception. Covariates in the models included mother’s age, education level, race/ethnicity, marital status, previous preterm history, plurality (number of previous live births), Kotelchuck index of prenatal care, pre-pregnancy BMI (underweight, normal, overweight, or obesity), drinking alcohol before pregnancy, and birth year. Groups with less than 20 mothers in the sample were not included in the figure (see **Appendix Table A3** for cell sizes of included and excluded groups).

Sensitivity analyses (**Appendix Tables S1** and **S2**) showed that the associations observed in the full sample also held in subgroups of young mothers and non-Hispanic white mothers who are most likely to vape, although some effect estimates were imprecise. In these subgroups, the excess risk of preterm births appeared to be limited to smokers and dual users who were also heavy smokers, and excess of risk of SGA is clearly seen among non-Hispanic white mothers (but less so among mothers <25 years of age) with either vaping or smoking.

## 4. DISCUSSION

### 4.1 Trend in the prevalence of e-cigarette use during pregnancy in 2016-2018

In a large national population-based sample of US women who recently had live births, we found that 3.6% of mothers used e-cigarettes within three months before conception and 1.1% of mothers vaped in late pregnancy. Previous studies on the prevalence of e-cigarette use during pregnancy are sparse. Among 1,071 pregnant women in the 2014-2017 National Health Interview Survey (NHIS), the prevalence of current e-cigarette use (“some days or every day”) was 3.6%.^14^ Among 864 participants in the Population Assessment of Tobacco and Health study (2013-2017), 2.8% of women reported vaping during pregnancy.^15^ However, none of these recent studies were based on nationally representative samples of pregnant women, accounted for dual use, or considered timing in pregnancy, and none of the studies accounted for errors in self-report.

We found that there was no observable change in vaping in our sample population of women with singleton live births from 2016 to 2018. This stable pattern over time is slightly different from the trend in the general adult population. The US nationally representative Behavioral Risk Factor Surveillance System reported that e-cigarette use prevalence in the general population was around 4.5% in 2016 and 2017, but increased by about 1% in 2018.^16^

### 4.2 Pregnancy outcomes associated with vaping in 2016-2018 PRAMS sample

We found that late-pregnancy sole vapers, especially if occasional, had similar risk of preterm birth as non-users. Although the relationship between tobacco smoking and higher risk of preterm birth has been well established, it is not fully understood which specific constituent of tobacco is responsible. Multiple mechanisms were proposed, including nicotine-induced vasoconstriction and carbon monoxide-induced fetal hypoxia.^17^ Use of e-cigarettes will eliminate exposure to smoking combustion products and certain other toxicants such as tobacco-specific nitrosamines,^18^ but not nicotine. Thus, the effect we see with frequent vaping has biological plausibility. In comparison to our earlier analyses of 2016 PRAMS data where we did not see evidence of vaping affecting risk of preterm birth,^8^ our current findings underscore the importance of considering the level of exposure, given that the excess risk from sole vaping, if any, appears to be limited to frequent users of e-cigarettes.

We observed that sole vapers and dual users in pregnancy had higher risk of SGA than non-users. This observation is consistent with existing literature, but somewhat at odds with our own analyses of sole vapers in 2016 PRAMS data that did not consider level of exposure.^8^ In utero nicotine exposure can increase the risk of fetal growth restriction by causing placental vasoconstriction and reducing fetal blood flow.^19,20^ However, the association with vaping was not consistent with exposure-response, weakening the argument for the importance of vaping alone in causing SGA, and again underscoring the importance of considering level of exposure.

Few studies have examined the relationship between exposure to cigarette smoke and vaping on pregnancy outcomes, and none to date have accounted for level of exposure. The National Academies of Sciences, Engineering, and Medicine published a consensus study in 2018 and concluded that there is insufficient evidence whether or not vaping affects pregnancy outcomes.^3^ One recent study by Kim and Oancea (2020) evaluated the association between vaping and pregnancy outcomes in the same data as we did.^9^ However, our work has methodological advantages that shed additional light on the problem. First, we retained about 95% of all women in the 2016-2018 PRAMS and our final study sample included 89,908 non-users, 8,164 sole smokers, and 723 sole vapers during pregnancy, allowing us to apply weights of the PRAMS sampling scheme. In contrast, Kim and Oancea discarded the majority of the initial PRAMS population due to inability to match on propensity scores, leaving only 28,939 non-users, 450 sole smokers, and 331 sole vapers in their final analytic dataset.^9^ This compromised the generalizability of their results and leaves them more vulnerable to chance findings due to loss of power. Additionally, the proportion of dual users is substantial: 63% of vapers in our sample also smoked traditional cigarettes concurrently, and in a study using 2014-2017 NHIS data,^14^ 39% of pregnant women who smoked also used e-cigarettes concurrently. However, Kim and Oancea excluded dual users and thereby did not investigate the full spectrum of smoking and vaping during pregnancy, limiting their conclusions to the minority of self-reported sole vapers. Exposure misclassification with unreported dual users in this small group is likely, given the well-known social desirability bias in disclosing smoking during pregnancy.

Control for confounding by race and age, due to concentration of vaping among younger non-Hispanic white mothers is a known concern for studies of pregnancy outcomes and vaping. We addressed this through stratified analysis and observed that such restrictions only strengthened conclusions about associations seen in full data, reducing concerns about bias from residual confounding and effect modification.

### 4.3 Subgroups of dual users by level of smoking and vaping

Previous studies typically assumed that dual users are a homogeneous group when evaluating their effects on pregnancy outcomes.^21^ In contrast, we acknowledged that dual users may include both habitual heavy smokers who occasionally vape and vapers who only occasionally smoke, leading to different risks of adverse pregnancy outcomes.

We observed that dual users had higher risks of SGA unless they were both light smokers and infrequent vapers. Nicotine is a known cause for fetal growth restriction,^19,20^ and our findings of higher SGA risks in dual user subgroups with higher presumed intake of nicotine (via self-titration theory)^22^ is biologically plausible. Our findings are also consistent with a study that compared levels of nicotine, cotinine, and tobacco-specific nitrosamines (TSNAs) in hair samples from a clinic-based sample of 76 mothers with singleton live births who reported current smoking and vaping in late pregnancy.^23^ That study reported levels of nicotine, cotinine, and TSNAs for pregnant dual users that were, on average, indistinguishable from those of smokers.

### 4.4 Limitations

Our study has several limitations. Maternal smoking and vaping were self-reported by mothers during a postpartum survey, which is bound to introduce misclassification of exposure, and such misclassification can be differential with respect to the outcomes.^24^ Although no validation study exists on the accuracy of self-reported maternal vaping before and during pregnancy in the PRAMS data, it is likely that self-report of vaping is more accurate than that of smoking since there is less stigma around vaping. We were unable to differentiate nicotine-containing versus nicotine-free e-cigarette used by pregnant women, a problem that may be especially severe for occasional vapers who may be less nicotine-dependent than frequent vapers (but we cannot be certain, especially since nicotine-free vapes may be more tolerated due to lack of saturation of nicotine craving). However, this may be a minor concern because the NHIS data showed that most adults reported vaping e-cigarettes that contain nicotine—91.2% of daily users, 88.2% of non-daily users.^25^ We are also not able to differentiate between closed-system e-cigarettes (disposable, non-refillable) that are under control of manufacturers and open-system e-cigarettes (refillable with e-liquids) which are more user-dependent.^26^ Open-system e-cigarette devices have been reported to be used for delivering ingredients such as different forms of marijuana and liquid tetrahydrocannabinol and, therefore, may confer different risks that can confound associations seen in our data.^27,28^

## 5. CONCLUSION

In a US nationally representative sample of women who had singleton live births in 2016-2018, we observed that dual users, regardless of their subcategories by smoking amounts and vaping frequencies, appear to have higher risk of SGA than non-users and conferred a similar risk as sole smokers. Although the effect estimates were imprecise, dual users who heavily smoked and sole frequent vapers may be at an increased risk of preterm births than non-users.

## Data Availability

The PRAMS Working Group and the Centers for Disease Control and Prevention collected and provided the data.

## Abbreviations

BMI: Body Mass Index
CDC: Centers for Disease Control and Prevention
NHIS: National Health Interview Survey
PRAMS: Pregnancy Risk Assessment Monitoring System
SGA: Small-For-Gestational-Age
TSNA: Tobacco Specific Nitrosamine

## Acknowledgement

The authors thank the PRAMS Working Group and the Centers for Disease Control and Prevention.

---

**Appendix Table A1.** Incidences and adjusted odds ratios (ORs) and 95% confidence interval (CI) for preterm births and small-for-gestational-age (SGA) births associated with dose of smoking and vaping in late pregnancy, **among non-Hispanic White mothers** in 2016-2018 PRAMS

| Dose of smoking and vaping in late pregnancy | Among non-Hispanic White mothers in 2016-2018 PRAMS<br>(Unweighted N 45,328) |  |  |  |  |
| --- | --- | --- | --- | --- | --- |
|  | Group<br>N | Preterm birth |  | SGA birth |  |
|  |  | Incidence % | Adjusted OR* | Incidence % | Adjusted OR* |
| **Non-users** | 40184 | 6.5 | Reference | 8.0 | Reference |
| **Sole occasional/frequent vapers** | 153 | 6.2 | 0.8 (0.4-1.5) | 14.3 | 1.7 (0.8-3.5) |
| **Dual users: light smoker and occasional/frequent vaper** | 255 | 6.6 | 1.0 (0.6-1.7) | 24.6 | 2.9 (1.8-4.8) |
| **Sole light smokers** | 2399 | 9.6 | 1.3 (1.0-1.6) | 19.0 | 2.3 (1.9-2.8) |
| **Dual users: heavy smoker and occasional/frequent vaper** | 249 | 9.1 | 1.5 (0.9-2.5) | 25.9 | 3.7 (2.3-6.0) |
| **Sole heavy smokers** | 2088 | 10.9 | 1.6 (1.2-2.0) | 23.2 | 3.2 (2.6-3.9) |

\*Adjusted for pre-pregnancy (in the 3 months before pregnancy) smoking/vaping (categorized as non-users, sole smokers, sole vapers, and dual users), mother's age, education level, marital status, previous preterm history, plurality (number of previous live births), Kotelchuck index of prenatal care, pre-pregnancy BMI (underweight, normal, overweight, or obesity), drinking alcohol before pregnancy, and birth year.

**Appendix Table A2.** Incidences and adjusted odds ratios (ORs) and 95% confidence interval (CI) for preterm births and small-for-gestational-age (SGA) births associated with dose of smoking and vaping in late pregnancy, **among young mothers (aged <=24 years at time of delivery)** in 2016-2018 PRAMS

| Dose of smoking and vaping in late pregnancy | Among mothers in 2016-2018 PRAMS<br>(Unweighted N 23,896) |  |  |  |  |
| --- | --- | --- | --- | --- | --- |
|  | Group N | Preterm birth |  | SGA |  |
|  |  | Incidence % | Adjusted OR | Incidence % | Adjusted OR |
| **Non-users** | 21084 | 7.7 | Reference | 10.8 | Reference |
| **Sole occasional/frequent vapers** | 99 | 8.5 | 1.0 (0.5-1.9) | 12.1 | 1.0 (0.4-2.2) |
| **Dual users: light smoker and occasional/frequent vaper** | 144 | 5.0 | 0.7 (0.3-1.7) | 14.9 | 1.1 (0.5-2.3) |
| **Sole light smokers** | 1652 | 10.6 | 1.5 (1.1-2.0) | 20.3 | 2.1 (1.6-2.7) |
| **Dual users: heavy smoker and occasional/frequent vaper** | 91 | 8.6 | 1.6 (0.7-3.9) | 19.8 | 1.8 (0.8-4.2) |
| **Sole heavy smokers** | 826 | 12.0 | 1.7 (1.2-2.4) | 22.7 | 2.6 (1.9-3.5) |

\*Adjusted for pre-pregnancy (in the 3 months before pregnancy) smoking/vaping (categorized as non-users, sole smokers, sole vapers, and dual users), race/ethnicity, education level, marital status, previous preterm history, plurality (number of previous live births), Kotelchuck index of prenatal care, pre-pregnancy BMI (underweight, normal, overweight, or obesity), drinking alcohol before pregnancy, and birth year.

**Appendix Table A3.** Unweighted frequency of mothers by pre-pregnancy and late-pregnancy smoking and/or vaping, among 2016-2018 PRAMS singleton births.

| **Dose of smoking and vaping in late pregnancy** | **Pre-pregnancy smoking/vaping** |  |  |  |  |
| --- | --- | --- | --- | --- | --- |
|  | **Non-users** | **Sole smokers** | **Sole Vapers** | **Dual users** | **Total** |
| **Non-users** | 79887 | 8093 | 915 | 1180 | 90075 |
| **Sole heavy smokers** | 19* | 2869 | 3* | 285 | 3176 |
| **Sole light smokers** | 35 | 4517 | 4* | 432 | 4988 |
| **Sole frequent vapers** | 0* | 33 | 33 | 65 | 131 |
| **Sole occasional vapers** | 0* | 24 | 12* | 72 | 108 |
| **Dual users: heavy smoker and frequent vaper** | 0* | 27 | 0* | 91 | 118 |
| **Dual users: light smoker and frequent vaper** | 0* | 30 | 0* | 133 | 163 |
| **Dual users: heavy smoker and occasional vaper** | 0* | 31 | 1* | 180 | 212 |
| **Dual users: light smoker and occasional vaper** | 1* | 38 | 0* | 191 | 230 |
| **Total** | 79942 | 15662 | 968 | 2629 | 99201 |

\* Groups with less than 20 mothers in the sample were not included in **Figures 1 and 2**.

